# An information-theoretic analysis of agrammatism in Broca’s aphasia

**DOI:** 10.1101/2023.04.23.23288999

**Authors:** Neguine Rezaii

## Abstract

Recently we proposed and tested a novel framework based on information theory about the features of agrammatism in patients with the nonfluent variant of primary progressive aphasia (nfvPPA). These features include short sentences, simplified structures, the omission of function words, reduced use of verbs, and increased use of heavy verbs. After distinguishing the syntactic and lexical features of agrammatism, we proposed that the lexical features are not defects but rather a compensatory response to structural deficits. We showed that although patients with nfvPPA have difficulty using complex structures, they use more informative words to facilitate the transfer of their intended message. In this work, we demonstrated the generalizability of our findings to patients with Broca’s aphasia who share similar neuroanatomical involvements with patients with nfvPPA. We found that patients with Broca’s aphasia use simpler syntactic structures as measured by higher syntax frequency. We also found that the use of simpler syntax predicts the use of lower-frequency words. Furthermore, our study showed that producing sentences of lower word frequency is likely achieved through the canonical features of agrammatism, i.e., higher proportions of content words over function words, nouns over verbs, and heavy verbs over light verbs.

## Introduction

Lesions to the left hemisphere’s inferior frontal gyrus are associated with a distinct style of language production known as agrammatism. These lesions may result from stroke or neurodegenerative disorders, such as primary progressive aphasia (PPA). Common symptoms of agrammatism include short, simplified sentences, the omission of function words, a decreased use of verbs relative to nouns, and increased use of heavy verbs than light verbs.^1–9^ The predominant account of the disease suggests that these symptoms result from a deficit in syntax processing.^6,10^ According to this account, the reduced use of function words and verbs in agrammatism can be attributed to their primary syntactic roles in a sentence, which often involve indicating grammatical relationships between content words.^6,10–16^ Under this account, however, it remains unclear why patients with agrammatism use more heavy verbs, have intact online access to the verb lexicon,^17,18^ and make minimal errors in using function words in sentence completion tasks.^19^

In a series of recent studies, first outlined by Rezaii et al. 2022,^20^ we proposed an alternative view of agrammatism based on information theory. We demonstrated that the lexical features of agrammatism are not deficits but rather a compensatory response to short and simple sentences to increase sentence information. A key requirement of this proposal is to distinguish between the syntactic and lexical abnormalities of agrammatism. The syntactic abnormalities would be short sentences and simplified structures. To measure the complexity of syntactic structures, we introduced syntax frequency, which measures the frequency of a given syntactic structure based on a large corpus of spoken language.^21,22^ Syntactic structures that are less common-- e.g., passive compared to active -- are more difficult to access, hence more complex. Using this metric, we showed that patients with the nonfluent variant of PPA (nfvPPA) use high-frequency syntax. On the other hand, the lexical abnormalities of agrammatism consist of increased use of content words over function words, nouns over verbs, and heavy verbs over light verbs. Our studies showed that patients with nfvPPA use lower-frequency words than healthy individuals. Crucially, we found a trade-off in the average frequency of words and the average frequency of syntax at the sentence level.^23^ This trade-off was true for both all words and content words. The syntax-lexicon trade-off thus predicts that if a sentence contains simple structures, its words will be more complex. These analyses were based on the language samples of participants as they described a picture to keep the overall message of language constant.

To show whether the lexical features of agrammatism arise as a result of simplified structures, we asked healthy speakers to describe the same picture using short sentences of one to two words. We found that in this constrained task, similar lexical features of agrammatism emerged in the language of healthy speakers. Constrained sentences contained more content than function words, more nouns than verbs, and more heavy verbs than light verbs. Interestingly, short sentences of healthy individuals contained more verbs in -*ing* form, a finding consistently reported in the language of patients with nonfluent aphasia. ^24–27^ These lexical features of short sentences resulted in a lower average word frequency when compared with unconstrained language. Lastly, we showed that using words of lower frequency in sentence-making results in higher information content of sentences. We measured information using lexical entropy and surprisal.^28,29^ The collective of our findings in nfvPPA suggests that the lexical profile of agrammatism is a compensatory response to short, simple sentences to increase sentence information.

In this study, we aim to test the generalizability of our findings to patients with Broca’s aphasia using language samples from the AphasiaBank.^30^ Previous research has shown considerable similarities between the language of patients with nfvPPA and Broca’s aphasia, likely due to their similar neuroanatomical defect in the left inferior frontal gyrus. ^31,32^ Given the similar language characteristics of the two groups, we hypothesize that patients with Broca’s aphasia 1) use simpler syntactic structures as measured by higher syntax frequency, 2) use lower frequency words as a compensatory response to simpler structures, and 3) show a trade-off between the frequency of words and syntactic structures which further highlights the lexical profile of agrammatism as a strategy to compensate for the loss of syntactic information.

## Methods

### Language samples

We obtained language samples from the AphasiaBank.^30^ We chose two topics for the picture description task, the cat rescue ^33^ (91 patients with Broca’s aphasia and 256 healthy controls) and the broken window stories ^34^ (86 patients with Broca’s aphasia and 271 healthy controls).

### Extracting language features

To extract various language features, we used Quantitext, a language toolbox developed in the MGH FTD Unit, to increase precision while reducing human labor.^35^ To determine the part of speech of words, the toolbox uses the automated Stanza Lexicalized Parser.^36^ We measured the proportion of <u>content words to all words</u> by dividing the number of content words by the number of all words in a sentence. Nouns, verbs (except *be, have*, and *do*), adjectives, and adverbs were considered content words. All other words were classified as function words. The proportion of <u>heavy verbs to all verbs</u> was measured by dividing the number of heavy verbs by the total number of verbs in a sentence. The following verbs were classified as light verbs: *‘be’, ‘go’, ‘take’, ‘come’, ‘make’, ‘get’, ‘give’*, and *‘have’* while excluding auxiliaries from this list.^37^ To measure <u>word frequency</u>, we used the Switchboard corpus,^38^ which consists of spontaneous telephone conversations averaging six minutes in length spoken by over 500 speakers of both sexes from a variety of dialects of American English. We use this corpus to estimate word frequency in spoken English, independently of the patient and control samples. The corpus contains 2,345,269 words. The word frequency of each sentence is calculated by taking the average log frequency of content words within that sentence. Therefore, in this manuscript, word frequency indicates the average log frequency of content words in a given sentence. To measure <u>syntax frequency</u>,^21^ we first parsed the sentences in the corpus using Stanza to extract headed syntactic rules. A headed syntactic rule is determined by the head and all its dependents in a dependency parse, whether they occur on the left or right. We applied this method on Switchboard, which resulted in 954,616 rules, and measured the syntax frequency of each sentence of participants by calculating the average log frequency of the syntactic rules of that sentence based on the Switchboard counts.

### Statistical analysis

For the statistical analyses of this study, we used the R software version 4.1.2. To compare language features at the sentence level across different groups, we used mixed-effects models with subject-specific random intercept via the lme4 package in R.^39^ We used independent t-tests to compare the features of agrammatism between patients and healthy controls.

## Results

### 1. The conventional description of the language of patients with agrammatism

Here, we first analyze the language of patients with Broca’s aphasia compared to healthy speakers to establish the existence of agrammatic features in their language. Following the convention of the field, these analyses were performed at the individual level (Figure 1).

**Figure 1.**
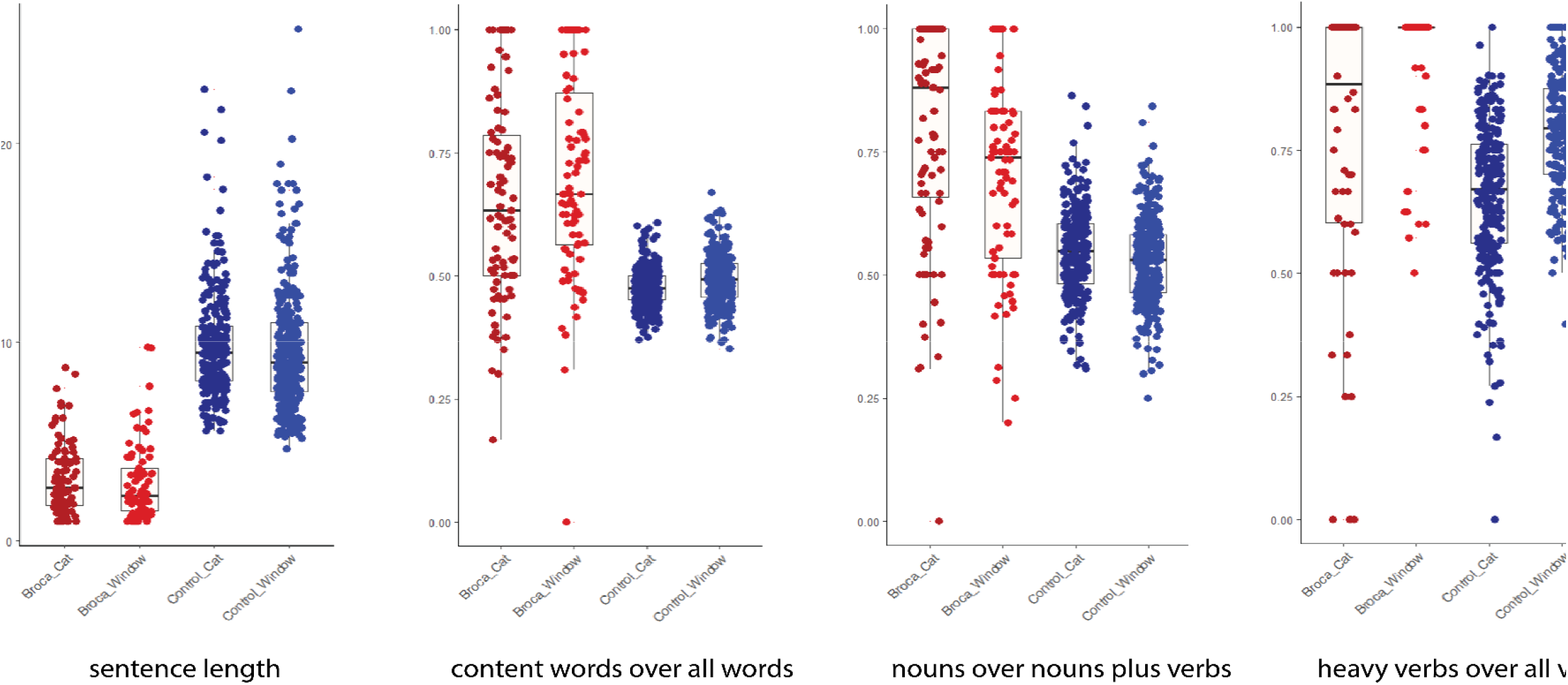
The box plots of various features of agrammatism in patients with Broca’s aphasia and healthy speakers across the cat and window stories

Cat story. Compared to the language of healthy controls, the sentences of patients with Broca’s aphasia were shorter (t(238.22) = -26.18, p < 0.001), with a higher proportion of content words to all words (t (92.484) = 8.373, p < 0.001), a higher proportion of nouns to the sum of nouns plus verbs (0.55) (t(100.77) = 10.138, p < 0.001) and a higher proportion of heavy verbs to all verbs (t(68.39) = 2.68, p = 0.009).

Window story. We found similar results in analyzing language samples obtained from the window story. Compared to the language of healthy controls, the sentences of patients with Broca’s aphasia were shorter (t(238.22) = -26.18, p < 0.001), with a higher proportion of content words to all words (t (88.57) = 9.38, p < 0.001), a higher proportion of nouns to the sum of nouns plus verbs (0.55) (t(94.74) = 7.83, p < 0.001) and a higher proportion of heavy verbs to all verbs (t(103.73) = 8.79, p < 0.001).

### 2. The information-theoretic analysis of language in Broca’s aphasia

For the information-theoretic approach, we analyzed language at the sentence level rather than the individual level to increase the sensitivity of analyses.

#### 2.1. Syntax frequency

We used a linear mixed-effects model to examine the relationship between *syntax frequency* and two predictor variables: *group* and *sentence length*. The model included random intercepts for subjects to account for individual variability in the response variable and to address the correlation among the repeated measures of syntax frequency for each subject. The results of the linear mixed-effects model for the cat story showed significant effects of both *group* (β= 0.279, t=2.663, p = 0.008) and *sentence length* (β= -0.047, t=-8.025, p < 0.001) on *syntax frequency* (Figure 2A). Specifically, the analysis revealed that patients with Broca’s aphasia produce syntactic structures of higher frequency than healthy controls. We used the same model on the data obtained from the window story and found significant effects of both *group* (β= 0.466, t=3.904, p < 0.001) and *sentence length* (β= -0.058, t=-8.025, p < 0.001) on *syntax frequency* (Figure 2B). The results suggest that regardless of the content of the picture across the two production tasks, patients with Broca’s aphasia use simpler syntax than healthy controls, as evident from their higher average syntax frequency in a given sentence.

**Figure 2.**
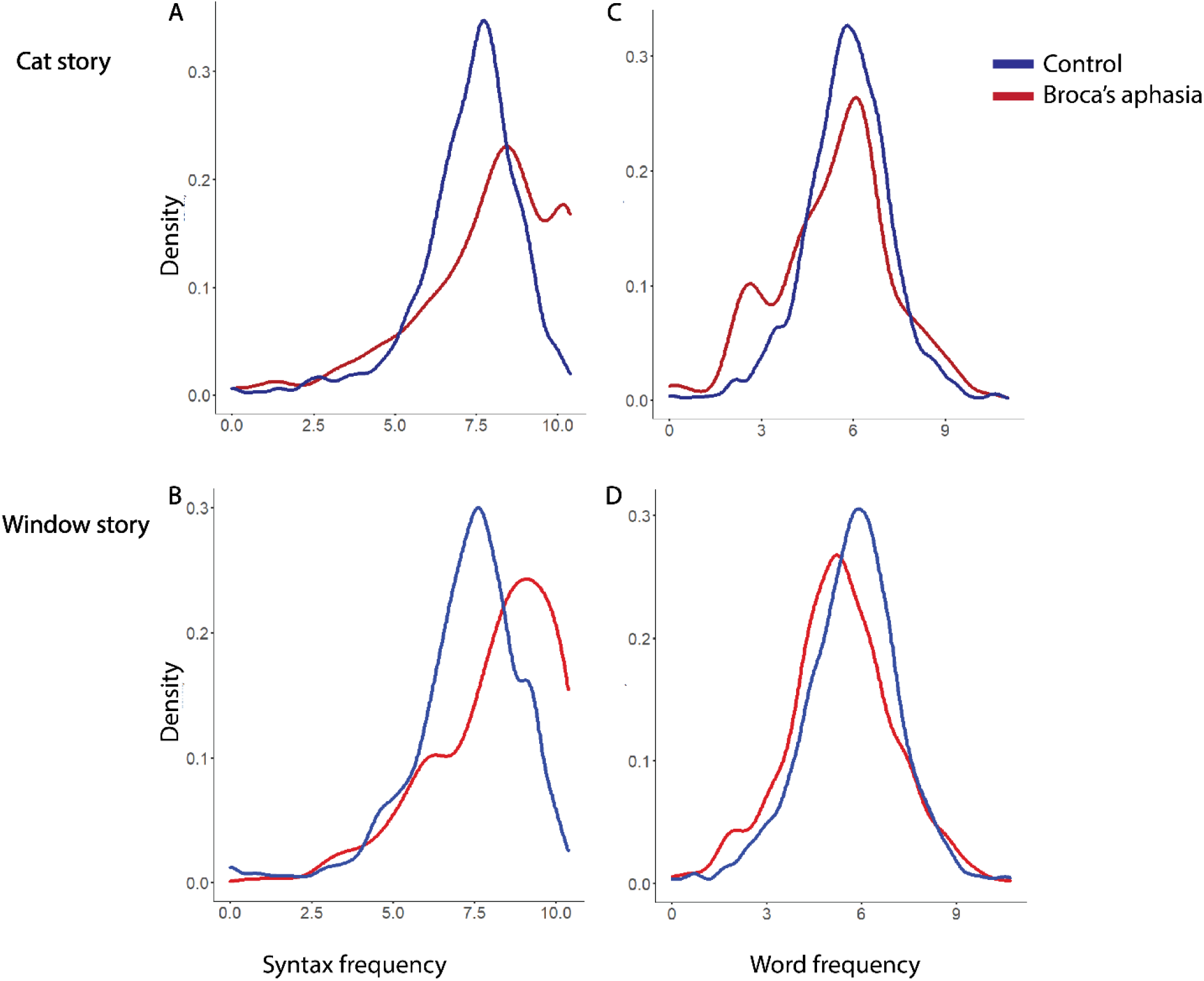
The density plots of syntax frequency and word frequency obtained from the speech samples of healthy speakers and patients with Broca’s aphasia describing the cat rescue and broken window pictures.

#### 2.2. Word frequency

Similarly, we compared the average frequency of content words between the two groups. We used a linear mixed-effects model to examine the relationship between the average log frequency of content words and two predictor variables: *group* and *sentence length* with random intercepts for *subjects*. The results of the linear mixed-effects model for the cat story showed significant effects of both *group* (β= - 0.254, t=-2.897, p = 0.004) and *sentence length* (β= 0.003, t= 5.140, p < 0.001) on *word frequency* (Figure 2C). We used the same model on the language samples obtained from the description of the window picture. Similarly, we found significant effects of both *group* (β= -0.182, t=-1.984, p = 0.048) and sentence length (β= 0.003, t=4.906, p < 0.001) on *word frequency* (Figure 2D). The results suggest that regardless of the content of the picture across the two production tasks, patients with Broca’s aphasia use lower-frequency words than healthy controls after controlling sentence length.

Here, we further show that the lower frequency of content words is associated with the canonical lexical features of agrammatism. We used a mixed effects model with random intercepts for *subjects* to predict word frequency from three predictor variables in the language of patients with Broca’s aphasia. We found that the lower frequency of words in a sentence could be predicted from a higher proportion of content words to all words (β= -2.453, t=-7.324, p < 0.001), a higher proportion of nouns over the sum of nouns plus verbs (β= -0.775, t=-2.878, p = 0.004), and a higher proportion of heavy verbs to all verbs (β= -2.073, t = -10.343, p < 0.001). These results suggest that the lexical profile of agrammatism reflects a strategy to lower word frequency, hence increasing sentence information.

#### 2.2. The syntax-lexicon trade-off

Here, we evaluated the trade-off between syntax frequency and word frequency at the sentence level. We used a linear mixed-effects model to examine the relationship between *syntax frequency* and two predictor variables, *word frequency* and *sentence length*, with random intercepts for *subjects* on the combined data from healthy individuals and patients with Broca’s aphasia. The results of the linear mixed-effects model for the cat story showed significant effects of both *word frequency* (β= -0.024, t=-13.115, p < 0.001) and *sentence length* (β= -0.005, t=-8.768, p < 0.001) on *syntax frequency*. Similarly, for the window story, we found significant effects of both *word frequency* (β= -0.289, t=-14.22, p < 0.001) and *sentence length* (β= -0.006, t=-11.13, p < 0.001) on *syntax frequency*.

We further examined the potential interaction effect of task (cat story vs. window story) with *word frequency* on *syntax frequency*. After combining data from the two picture description tasks, we found that the interaction term, *word frequency* : *task*, was not a significant predictor of *syntax frequency* (β= -0.004, t=-1.629, p = 0.103), suggesting that the syntax-lexicon trade-off was independent of the content of the picture.

Further, we examined the potential interaction effect of *group* (Broca vs. healthy controls) with *word frequency* on *syntax frequency* and found that the interaction term, *word frequency* : *group*, was not a significant predictor of *syntax frequency* (β= 0.001, t=-0.136, p = 0.892). This finding suggests that the trade-off is independent of the group because healthy controls and patients with Broca’s aphasia show similar trade-offs (Figure 3).

**Figure 3.**
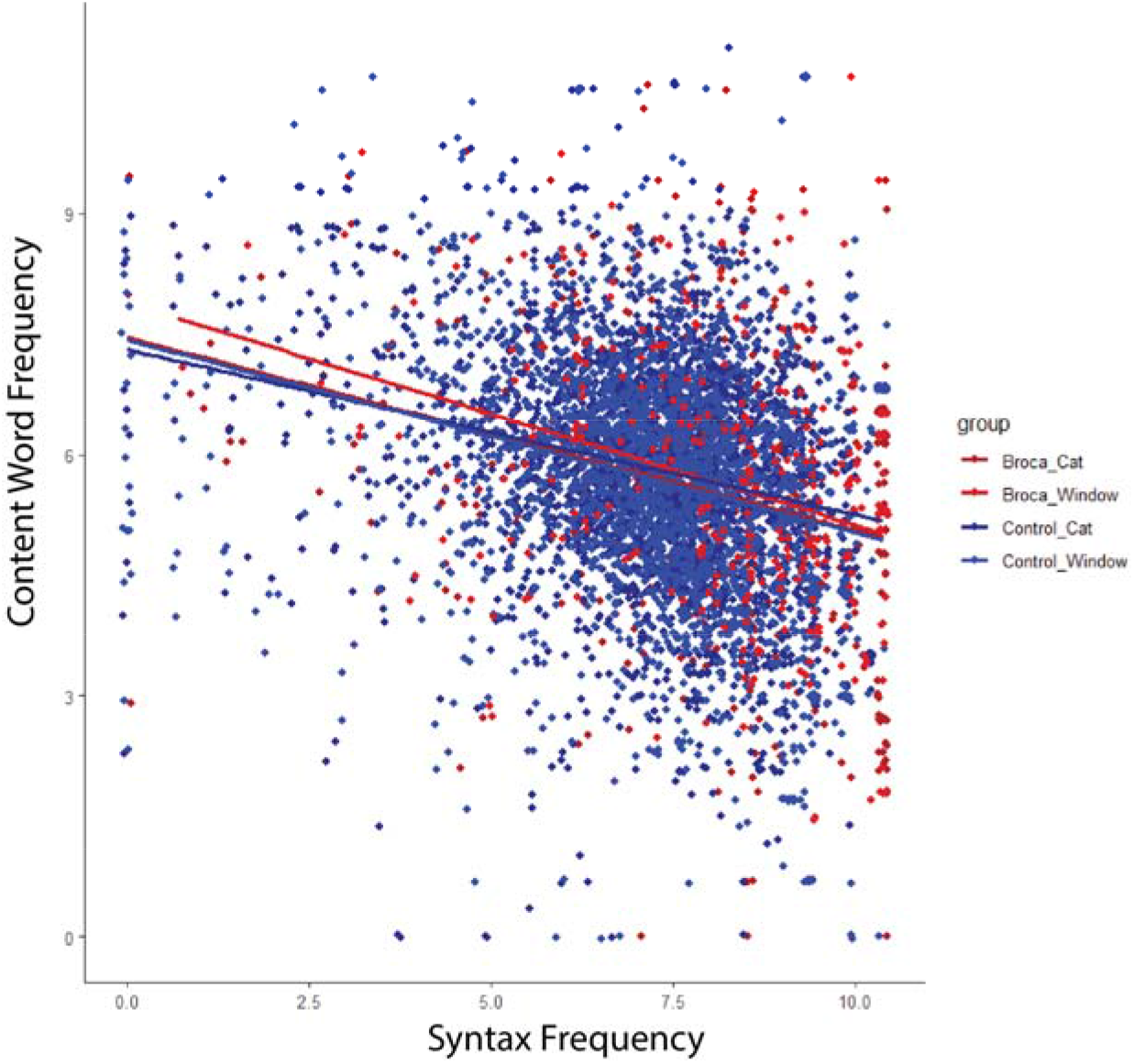
The syntax-lexical trade-off in the language of patients with Broca’s aphasia (in red) and healthy speakers (in blue) obtained from the description of cat and window stories.

## Discussion

In this work, we provided evidence synergistic with our information-theoretic proposal about agrammatism. Previously, we showed that the lexical profile of patients with nfvPPA is not a defect but rather a compensatory response to facilitate the transfer of their intended message. Specifically, patients with nfvPPA use more informative words to compensate for the loss of information that was supposed to be carried by syntactic structures.^28,29^

This study extended the generalizability of our previous findings on nfvPPA to patients with Broca’s aphasia. This generalizability was expected, although not shown, given the overlapping neuroanatomical involvement between nfvPPA and Broca’s aphasia. We first found that syntax frequency is an effective metric to capture structural deficits in patients with Broca’s aphasia, as these patients produced high-frequency syntactic structures. We then found that patients with Broca’s aphasia use lower-frequency content words compared to healthy individuals after controlling for sentence length. Producing sentences of lower-frequency words is likely achieved through the canonical lexical profile of agrammatism. The lexical profile that significantly correlated with a lower word frequency included higher proportions of content over function words, nouns over verbs, and heavy verbs over light verbs. Lastly, we found a syntax-lexicon trade-off in the language of patients with Broca’s aphasia as well as healthy individuals. The collective findings suggest that individuals with nonfluent aphasia, including those with Broca’s aphasia and nfvPPA, tend to select specific types of words to enhance the information content of their sentences. This compensation mechanism appears to be a response to their difficulty in generating long and complex sentences.

The results of this study were based on the description of two different pictures. Picture description tasks have the advantage of controlling the total amount of information needed to convey. We found similar results independent of the topic of the two pictures. Of note, the two pictures used in this study (cat rescue and broken window) were different from the picture we used in our studies on nfvPPA patients (picnic scene)^40^ despite similar results. Future research is needed to investigate whether the information-theoretic features described in this work continue to exist in casual, everyday conversations. Furthermore, we conducted our analyses of language at the sentence level rather than the individual level. Previously, we showed that healthy speakers have the ability to shift between the use of complex syntax and complex words from sentence to sentence. For example, they might use simple words and complex syntax in one sentence while they adopt the opposite strategy in another. Therefore, analysis of language at the individual level that averages the syntax frequency and word frequency of all sentences would be insensitive to the normal variability used by healthy speakers.

From this work, it remains unclear what the origin of the syntactic impairment in patients with nonfluent aphasia is. For example, it is unclear whether it is the inability to make complex structures that result in shorter sentences or it is the inability to make long sentences that forces nonfluent patients to use simple syntax. Previous theories of compensation attributed the difficulty of making long, complex sentences to the cost of articulation.^41^ However, we recently showed a high degree of similarity between the speech and writing of patients with nfvPPA^28,42^ confirming previous studies showing similar patterns in patients with Broca’s aphasia.^43^ The similarity between writing and speaking excludes the cost of articulation to be the origin of the structural deficits in patients with nonfluent aphasia. Future work is required to delineate the functional locus of deficit as the bottleneck in producing syntactically complex structures.

## Data Availability

The data is available through the AphasiaBank

## Notes

### Competing Interest Statement

The authors have declared no competing interest.

### Funding Statement

Nothing to declare

